# Structural and EEG motor networks distinguish level of motor impairment after stroke

**DOI:** 10.1101/2025.10.08.25337630

**Authors:** Zhibin Zhou, Anirudh Wodeyar, Steven C. Cramer, Ramesh Srinivasan

## Abstract

The primary objective of this research is to elucidate brain connectivity patterns that are associated with motor impairment in individuals recovering from stroke. Here we estimate individual patient (1) structural connectivity using stroke lesions modeled from MRI and (2) brain functional connectivity from EEG using graphical modeling informed by the structural connectivity of individual patients. Our analyses concentrated on the brainstem and three motor-related cortical areas in both hemispheres: precentral cortex (primary motor cortex), caudal middle frontal cortex (dorsal premotor cortex), and superior frontal cortex (incorporating the supplementary motor cortex).

Network analysis metrics were applied to structural and functional data, including degree centrality, average efficiency, and betweenness centrality. These measures were used to contrast motor networks in patients divided into two groups based on their upper extremity motor deficits. We found that structural disconnection of the brainstem from both dorsal premotor cortex and primary motor cortex indicated the highest probability of low motor status. All structural connectivity metrics of the 3 ipsilesional motor areas were higher in the higher motor status group. We integrated structural connectome with source localized EEG data to estimate the network analysis metrics. The functional connectivity revealed a notable pattern similar to the structural connectivity in the δ band (1-3 Hz) within ipsilesional motor areas. In addition, contralesional motor areas showed enhanced connectivity in the β band (14-29 Hz) in patients with higher motor status, possibly as a result of compensatory neural plasticity of the contralesional homologues.

We also introduced a novel metric, motor betweenness centrality, to quantify the role of non-motor regions in rerouting motor-related communication for both structural and functional network analyses. Almost all new hubs for routing the communication with motor-related areas identified in the structural connectome were associated with lower motor status, suggesting network reorganization in patients with greater impairment is inefficient. However, EEG-based routing analyses further revealed that high motor status patients engaged nearby cortical areas, while low motor status patients rerouted signals through more distant and less efficient regions. EEG connectivity metrics can potentially be measured at multiple points during treatment and may reflect functional plasticity.

In summary, the structural and EEG-based functional network properties—degree centrality, efficiency, and betweenness centrality—reliably identified key motor ROIs predictive of motor status. Motor betweenness centrality captured plasticity-related rerouting outside primary motor regions. Delta and beta band EEG networks were most informative of motor recovery. There was notable overlap between the structural and functional connectivity findings. In particular, ipsilesional motor areas, especially M1 and PMd, were consistently identified as critical hubs in both modalities. Structural disconnection of these areas predicted poor motor status, while higher EEG connectivity in these regions in δ and β bands predicted better outcomes. These findings underscore the clinical potential of combining multimodal network measures to monitor, predict, and guide recovery in stroke rehabilitation.

## Introduction

Evaluation of behavioral status after stroke is crucial for measuring specific impairments and functional disabilities and for treatment planning. The motor system is affected in most patients with stroke (Rathore et al., 2002). Tools for measuring motor status using behavioral evaluation are widely used in clinical and research settings (Gladstone, Danells, Black 2002; Rathore et al., 2002, and See et al., 2013). Such assessments are instrumental in creating personalized rehabilitation plans. Conducting these evaluations early and accurately can yield valuable prognostic insights, offering predictions about the trajectory of a patient’s recovery. This prognostic information is essential for healthcare providers and patients alike, as it helps establish realistic goals and expectations for the rehabilitation process (Veerbeek et al. 2011).

Stroke injury and recovery are direct consequences of structural and functional modifications in the brain. Patients who experience a stroke often sustain damage to the white matter tracts, altering both structural and functional connectivity. However, the heterogeneity of lesion location and extent in stroke presents significant challenges in developing models for predicting post-stroke recovery directly from structural changes due to injury. Stroke is often considered a ‘circuitopathy’(Campos et al. 2023; Cassidy et al., 2022; Meder and Siebner,2018), where even a single disconnection can alter brain function and behavior. Structural neuroimaging measurements that reflect corticospinal tract injury have been well documented in the literature as potential biomarkers for post-stroke motor recovery (Cassidy et al., 2018; Lin et al., 2019). Similar studies of structural connectivity, such as those focusing on the connectome’s role in motor status recovery, reveal that disrupted structural and functional connectivity in large-scale neural networks contributes to neurological deficits and recovery potential after stroke (Lim and Kang 2015). Structural connectivity between brain regions remote from infarct lesions demonstrates correlations with somatosensory and motor outcomes, emphasizing the importance of these distant neural pathways in functional recovery (Koh et al. 2021). Additionally, longitudinal changes in white matter microstructure reflect both degenerative and adaptive reorganization processes, which are thought to influence motor recovery outcomes following stroke (Koch et al. 2021).

Connectome-based predictive modeling has successfully identified neuronal networks underlying motor recovery, providing valuable insights into individualized therapeutic strategies for post-stroke patients(Olafson et al. 2021; Koch et al. 2021) and response to rehabilitation therapies beyond what can be learned from measures of neural injury (Burke Quinlan et al., 2015). Moreover, research highlights the critical role of interhemispheric structural connectivity, such as the connection between primary motor areas (M1-M1), in enabling complex motor control and facilitating recovery(Paul et al. 2023). Structural connectivity changes in the motor execution network have also been associated with functional recovery during post-stroke rehabilitation(Wanni Arachchige et al. 2021). In this paper, we examine the relationship between damage to the structural connectome, estimated from lesion location and extent, and level of motor deficits after stroke.

The functional organization of the brain does not have a 1:1 correspondence with brain anatomy (Griffis et al., 2019; Wodeyar et al 2020, and Reid et al., 2019), and so functional data provide an additional dimension of insight into post-stroke recovery (Cassidy, Mark, and Cramer 2022). Furthermore, while structural connectivity is generally considered less plastic than functional connectivity, lesion-related structural alterations are expected to remain relatively stable over time, functional connectivity evolves substantially with stroke recovery and may reflect functional plasticity (Wanni Arachchige et al., 2021). Studies using fMRI have shown how functional connectivity strength increases between relevant brain regions over time, aligning with motor recovery. For example, increases in functional connectivity within the ipsilesional motor cortex and improvements in interhemispheric connections often predict motor function gains (Ismail, Yahya, and Manan 2024).

Electroencephalography (EEG) provides a noninvasive, cost-effective bedside method for assessing functional connectivity and neural plasticity during stroke recovery (Keser et al., 2022). Functional connectivity in EEG can be measured by the coherence of neural oscillations, which measures the consistency of phase and amplitude differences over time, and may reflect communication between spatially distinct neural populations mediated by the white-matter fibers of the brain (Fries 2005; Nunez and Srinivasan 2006; and Schneider et al., 2021). These measures serve as valuable biomarkers of cortical function and plasticity (Wu et al. 2015). EEG coherence between the ipsilesional primary motor cortex and various locations in both the ipsilesional and contralesional hemispheres was found to explain 62% of the variance in motor recovery, outperforming EEG power, baseline motor status, and corticospinal tract injury (Cassidy et al. 2021). Low-frequency oscillations (1-3 Hz) have been identified as biomarkers of injury and recovery after stroke (Cassidy et al. 2020).

EEG measures of functional connectivity of the motor system, particularly between the ipsilesional primary motor cortex (M1) and ipsilesional dorsal premotor cortex (PMd), are strongly associated with motor deficits and their improvement through therapy after stroke(Kantak et al., 2012). However, functional connectivity is not directly comparable to the structural connectivity since it can emerge from both direct and indirect structural connections, also known as common input effects (Wodeyar and Srinivasan 2022). We have developed an approach to integrate structural connectivity with functional data to estimate a graphical model of functional connectivity of fMRI (Wodeyar et al 2020) and EEG/MEG (Wodeyar and Srinivasan 2022). Using this approach we have modeled how damage to structural connectivity modulates fMRI functional connectivity in stroke patients (Wodeyar et al 2020). In EEG/MEG we incorporate the structural connectome into a graphical model to estimate partial coherence (Wodeyar and Srinivasan 2022).

Partial coherence is the estimate of the direct relationship between two brain areas after removing the influence of all other brain areas, thereby minimizing the effects of common inputs and volume conduction (Wodeyar and Srinivasan 2022).

This paper uses individual patient models of structural connectivity and graphical modeling of EEG functional connectivity to explore how a patient’s level of motor deficits after stroke is related to structural injuries and mediated by functional connectivity. Our focus is on the connectivity of the brainstem with specific motor-related cerebral cortical areas: precentral gyrus (M1), superior frontal gyrus (SFG, which includes supplementary motor area - SMA), and caudal middle frontal gyrus (PMd for the upper limbs). Based on previous studies, we hypothesized that the delta (δ) and beta (β) frequency components of EEG will reflect aspects of motor network communication that are most related to motor status. We introduce a novel approach to characterizing plasticity of whole-brain connections of the motor system following brain injury. Our results demonstrate how both structural and functional network properties reliably capture motor status after stroke.

## Methods

### Data Acquisition

This study was conducted on de-identified datasets and all procedures adhered to the principles of the Declaration of Helsinki. The study protocol received approval from the Institutional Review Board. We analyzed structural MRI and EEG data from 57 patients diagnosed with stroke. MRI images for all patients were obtained at the time of admission to the acute care hospital, while EEG was collected at various time points thereafter (see Figure 2). Each patient’s motor status was assessed at the time EEG data were collected. Since EEG recordings spanned patients from the acute to chronic stages of recovery, this heterogeneity provided an opportunity to test the robustness of our network measures across different biological states post-stroke. Structural connectivity captured relatively stable lesion-related disruptions, while EEG functional connectivity reflected adaptive plasticity observable at multiple recovery stages. Neuroimaging was acquired on a 3.0 T Philips Achieva (Best, the Netherlands) scanner.

Anatomical imaging included a high-resolution T1-weighted scan that utilized a three-dimensional magnetization-prepared rapid gradient echo sequence (repetition time (TR)=8.1 ms, echo time (TE)=3.7 ms, 150 slices) and a T2-weighted FLAIR scan (TR= 9,000 ms, TE=120 ms, 33 slices).

EEG data were recorded at 256 channel electrodes (Electrical Geodesics, Inc.) for 3 minutes at rest, with eyes open and fixation on a red cross displayed on a computer monitor to minimize eye movements. Artifacts were removed from the data using entirely automated procedures: (1) 48 channels located on the face and neck were discarded, as most of that data was artifact, leaving 208 channels on scalp and forehead in the analysis; all further steps only use these 208 channels. (2) Bad electrodes had a variance above a specific threshold (> 2.5 standard deviation). (3) The data were filtered between 0.25 Hz (stopband 0.01 Hz) and 50 Hz (stopband 51 Hz) with a Butterworth filter with 6 dB passband ripple, and 20 dB stopband attenuation. (4) ICA (independent component analysis) was used to decompose the data using the FastICA algorithm. (5) Independent components having correlation coefficients higher than 0.4 with signals from electrodes closest to the eyes were considered eye blink artifacts and were removed. (5) Independent components highly correlated with signals from bad electrodes were also removed. (6) The data were then reconstructed without artifact components at the original 208 electrodes.

Motor deficits were assessed using the Fugl-Meyer Assessment of the Upper Extremity (FMA-UE) Scale of Motor Impairment (hereafter called FM) (See et al., 2013).

### Lesion Masks

Lesion masks were drawn on T1-weighted brain MRIs, informed by the corresponding T2 FLAIR images. Masks were generated using techniques (Cassidy et al. 2018; Riley et al. 2011) for which reliability has been described previously (Burke et al. 2014). For one subject who was unable to complete an MRI scan, the lesion mask was obtained from their corresponding head CT scan. Infarct masks were binarized and spatially transformed to MNI space. For consistency, unilateral lesions on the right were flipped about the midline so that most lesions were on the left side of the brain for analysis purposes. Correspondingly, the EEG data for those patients were also spatially flipped to align with the lesion orientation. For consistency, the left hemisphere is referred to as the ipsilesional side (denoted as “I” in figures and tables), while the right hemisphere is referred to as the contralesional side (denoted as “C”).

### Patient Stratification

Out of the 57 stroke patients in our dataset, 7 had lesions localized exclusively to the brainstem. In our network model, connections are represented as undirected edges between cortical regions of interest (ROIs). Because the brainstem is represented by only a single ROI, including it introduces artificial interhemispheric connections, which may distort both structural and functional connectivity estimates.

To address this, we conducted separate analyses:

- For characterizing lesion-induced structural disconnections involving the brainstem, we included all 57 patients and examined structural connections between the brainstem and cortical ROIs.
- For all other structural network analyses—including interhemispheric comparisons and centrality metrics—we excluded brainstem ROIs and focused on the remaining 50 patients whose lesions involved supratentorial regions. However, since EEG source reconstruction was limited to cortical ROIs, all 57 patients were included in the EEG analyses.

We divided the patients into 2 groups by their FM scores. Patients with a FM score of ≥ 42 are placed in the high level group (H). Patients with an FM score of 42 are placed in the low level group (L) (Woytowicz et al., 2017). There are 28 and 29 patients respectively in the H and L groups out of all 57 patients. Out of the 50 patients, there are 24 and 26 patients in the H and L groups respectively.

### Structural connectivity

#### Individualized Reduced Structural Connectome

A full intact normative template of brain structural connectome was computed by combination of using a probabilistic atlas based on Lausanne parcellation, dividing the brain into 463 ROIs (Cammoun et al. 2012) and streamlines (white matter tracks) generated from tractography analysis of a group average template of DTI constructed from the HCP 842 data set (Yeh et al. 2018; Van Essen et al. 2013). A total of 133,815 streamlines were obtained using the FMRIB Software Library (FSL) (Jenkinson et al. 2012; Woolrich et al. 2009; Smith et al. 2004) which were transformed to the MNI152 space. The streamlines were approximated with adapted code from the along-tract-stats toolbox (Colby et al. 2012). We mapped the connections of these streamlines to the 463 ROIs (Daducci et al. 2012) from Lausanne parcellation in the same volumetric space.

There are 448 cortical ROIs and 15 subcortical ROIs generated by easy_lausanne toolbox (Daducci et al. 2012). Pairs of ROIs identified as being connected by the intact streamlines were used to construct the structural connectome.

The structural connectivity of each patient was modeled using a virtual tractography method (Kuceyeki et al. 2013; Wodeyar 2019; Wodeyar and Srinivasan 2022).

Individual patient’s reduced structural connectomes were constructed by intersecting the location of the volumetric lesion with the streamlines to determine which regions were disconnected. Pairs of ROIs identified as being connected by the remaining streamlines not damaged by the stroke lesion were used to construct the matrix of the individualized reduced structure connectome for each patient. As illustrated in Figure 1B, this procedure generates three complementary matrices: the full structural connectome derived from tractography, the subset of connections intersected by the lesion, and the reduced structural connectome capturing the preserved pathways.

**FIGURE 1.**
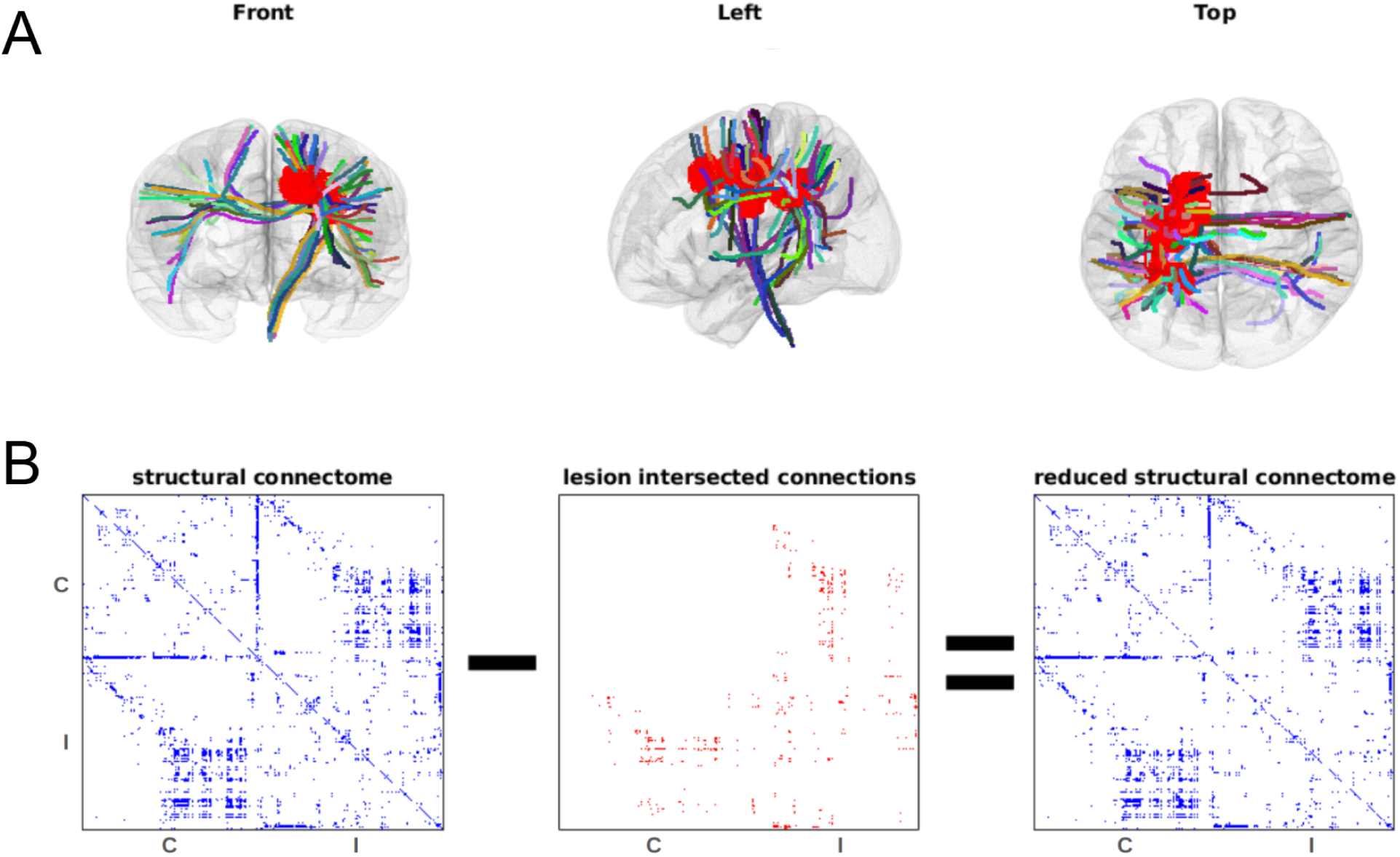
Reduced Structural Connectome: A. The stroke lesion mask is shown in red in a patient with a unilateral hemispheric infarct. Streamlines that intersect the lesion are shown by the colored lines. B. The 3 matrices below from the left are the structural connectome, the lesion intersected connections, and the reduced structural connectome for the 463 ROIs of the Lausanne parcellation (scale 250). We use the binarized structural connectome, with any non-zero edge shown in blue.

Comparing these side by side emphasizes how stroke lesions selectively disrupt certain connections while leaving others intact, thereby clarifying the residual network architecture. For the purposes of our analyses, ROIs that were only partially disconnected were still considered connected, since streamline counts are not a reliable measure of fiber integrity, and even a small number of surviving streamlines may reflect a meaningful structural connection.

#### Brainstem connectivity of the motor system

We used the structural connectome to make a model of the connectivity of the brainstem to the contralesional and ipsilesional motor areas. For each ROI in the motor system, we computed the probability of being in the low FM score group given that the ROI is disconnected from the brainstem *P*(*L*|*D*):

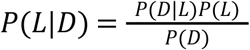

In the above formula, P(L) is the probability of being in the low FM score group, P(D) is the probability of the ROI being structurally disconnected to the brainstem, and *P*(*D*|*L*) stands for the probability of an ROI being structurally disconnected given the patient is in the low FM score group.

All network measurements from stroke patients were normalized to the healthy control brain. To facilitate interpreting the contralesional hemisphere, we examined the outputs of our normalized degree centrality after removing the interhemispheric connection in the networks. We confirmed that, for all patients with unilateral stroke lesions, the degree centrality on the contralesional side is identical to a control brain without interhemispheric connections. Thus, we can interpret contralesional changes in network properties in patients as entirely due to disconnection with the ipsilesional hemisphere.

### EEG functional connectivity

#### Constructing the head model for source localization

The source model was created in MNE (an open-source Python package) (Gramfort et al. 2013). The locations of the 256 EEG sensors were spatially aligned with the digitized scalp landmarks using fiducials from the standardized Freesurfer fsaverage (Fischl 2012) surfaces with MNE’s co-registration process. The boundary element method (BEM) was used to create a forward model of the head geometry using the standardized *fsaverage* surfaces. The conductivity values for each of the three layers (inner skull, outer skull, and skin) for EEG are 0.3, 0.0075, and 0.3. This procedure created 5,124 source locations with free orientations, which were then converted to surface-based source orientations with each dipole having a fixed direction perpendicular to the brain surface. It produced a Lead Field Matrix (LFM) (*G*) of dimension 256 * 5,124 (256 sensors by 5,124 dipoles). The 5,124 dipole positions were evenly spaced and constrained to the brain surface (using icosahedron subdivision grade 4). This EEG leadfield matrix was used to generate the inverse solution, removing the 48 channels located on the face and neck, which were heavily contaminated with artifacts. We validated the model by confirming that the reconstructed EEG from the source model has (across channels and patients) an average correlation 0.929 with the original EEG signals.

#### Source data aggregation in each ROI

Each of the 5,124 sources was labeled with one of the 448 cortical regions of the Lausanne Parcellation (scale 250) of the fsaverage brain (Cammoun et al. 2012; Daducci et al. 2012). We aggregated the source-localized data of the dipoles within each ROI using PCA (principal component analysis). The first principal component accounted for 86.2% percent of variance on average in each ROIs from all patients. By representing the time series of each ROI using only the first principal component, we reduced the number of EEG source time series from 5,124 to 448 (only including the ROIs of cortical brain regions).

#### Estimating partial coherence between sources

Each source time series was bandpass filtered into six frequency bands δ (1-3 Hz), θ (3.5-6.5 Hz), α (7-10 Hz), µ (10.5-13.5 Hz), β_1_ (14-20 Hz), and β_2_ (21-29 Hz), using Matlab’s elliptical filter design with stopband attenuation set to 60 dB, passband ripple set to 0.2 and filter order of 20. The filtered signals in each band were Hilbert transformed to obtain analytical signals *A*. The analytical signal *A* captures the instantaneous amplitude and phase of the signals.

We used the analytical signals to calculate cross-spectral density between each ROI pair. This is equivalent, up to a linear transformation, to estimating the cross-spectral density using a windowed Fourier transform (Lepage et al., 2013). The cross-spectral density can be normalized by the product of the power (squared amplitude) in each ROI to estimate coherence, a widely used measure of functional connectivity in EEG (Nunez and Srinivasan 2006). Coherence estimates have limitations due to volume conduction (Srinivasan et al. 2007) and the effects of common inputs (Wodeyar and Srinivasan 2022). To address these concerns, we estimated partial coherence between ROIs using a graphical model of functional connectivity ((Wodeyar and Srinivasan 2022)) constrained by the structural connectome. Partial coherence measures direct functional connectivity between two ROIs after removing the influence of all other ROIs (i.e., indirect connections) which also minimizes volume conduction effects. Our method utilizes the Graphical Lasso (GL) (Friedman, Hastie, and Tibshirani 2008) method using the QUIC algorithm (Hsieh et al. 2011). We estimate a sparse model of direct connections between ROIs which can account for the measured cross-spectral density. In this procedure, we can incorporate our knowledge of each patient’s reduced structural connectome by optimizing two penalty terms (Wodeyar and Srinivasan 2022):

1. penalization for the patient-specific reduced structural connectome edges - λ_*c*_ and
2. penalization for all other edges - λ_*o*_. We split the data into two ensembles, and used cross-validation to select the penalty values. In all patients and frequency bands, the value of λ_*c*_ selected was always smaller than λ_*o*_ indicating that the structural connectome provided useful information to fit the model of cross spectral density.

### Characterizing structural and functional networks using graph theory

For each patient, we created a structural network graph and 6 functional network graphs, one in each EEG frequency bands (δ, θ, α, µ, β_1_, β_2_). The structural network graph *G* had 463 cortical and subcortical nodes (corresponding to ROI) and edges *E*_*est*_ = *G*_*ij*_, *i*, *j*∈*N* obtained from the reduced structure connectome. For each of the EEG frequency bands we had 448 cortical nodes with edges *E*_*est*_ = 1, *ifPC*_*ij*_ > 0, *i*, *j*∈*N* derived from the non-zero values in the partial coherence (*PC*) estimates.

The network’s edges are undirected and unweighted. We estimate the following network metrics using the Networkx Python package (Hagberg, Schult, and Swart 2008).

1. Degree centrality reflects the number of direct structural/functional connections of an ROI. It is the fraction of all other nodes (ROIs) connected to each node. In our analysis, we focus on the degree centrality of ROIs with the 3 motor-related brain areas (M1, PMd, and SFG).
2. Average efficiency is a metric that tries to express how efficiently an ROI is can communicate with other ROIs in the brain. It is the inverse of the shortest path length from each node to all other reachable nodes (note that this is modified from the NetwrokX implementation). A direct connection is the shortest possible path with an efficiency of 1, while two disconnected nodes have an efficiency of 0. For each node, we average the efficiency over all 447 possible connections to other nodes. In our average efficiency analysis, we focus on only the ROIs of the 3 motor-related cortical areas of interest to evaluate the efficiency with which each node in the motor system communicates with the rest of the brain.
3. Betweenness centrality of each ROI is the fraction of the number of shortest paths of the entire network that go through or end at that ROI. An ROI with high betweenness centrality could gate considerable activity within the brain network. We focused on betweenness centrality of nodes in the 3 motor-related areas to measure information flow through motor-related brain areas. In the patients studied here, this measure is expected to be sensitive to potential motor status related communication disruption.
4. Motor betweenness centrality is the fraction of the number of shortest paths starting from the motor-related areas through non-motor ROIs. Specifically, for any non-motor ROI A, it is calculated as the number of shortest paths from motor ROIs to all other ROIs that pass through A, divided by the total number of shortest paths from motor ROIs to all non-motor ROIs. Thus, this measures the importance of a non-motor ROI in relaying/receiving/giving motor signals on the existing routes of the damaged structural network or the (potentially) newly formed routes of the functional network during the motor recovery post-stroke.

For motor betweenness centrality analysis, we analyze all the ROIs outside of the 3 motor-related brain areas.

For normalize, all network metrics derived from each patient’s reduced structural connectome were expressed as a percentage of the corresponding values from the intact structural connectome. For degree centrality, average efficiency, and betweenness centrality, we focused on examining only the ROIs of the 3 motor-related areas. For motor betweenness centrality, we explored all other brain regions outside the motor-related areas.

### Statistical tests

Comparison of network measurements between the high and low-motor level groups using Student’s t-test is very sensitive to outliers. As a result, we used a statistical method involving permutation simulation with a preset significance level α. The permutation method is a robust approach to determining the significance of an observed effect. This method is particularly useful in scenarios where traditional parametric assumptions (e.g., t-test) may not hold, or when the exact distribution of the test statistic under the null hypothesis is unknown. For the current permutation method, after the network statistics were computed, we calculated the ratio of their real group means H/L and performed a permutation test by randomly shuffling the group labels. The random resampling (permutation) was done 100,000 times for each network measurement to obtain a null distribution of the simulated H/L values for each ROI. In the case of structural connectivity, as we have no reason to believe losing connections will lead to higher motor status, we only tested H > L (α=0.05). For EEG measures, which potentially reflect functional plasticity, we tested both directions (α=0.05).

## Results

### Demographics and Anatomy

Patients’ characteristics are listed in Table 1. Infarct volume, age, and the number of days post-stroke showed no correlation with their motor status (FM scores; Figure 2).

**Table 1:** Patients’ characteristics.

| | Mean ± SD |
| --- | --- |
| Total number of patients | 57 |
| Age (year) | 56 ± 14 |
| Lesion size (mm<sup>3</sup>) | 44296 ± 81156 |
| Time post-stroke (days) | 376 ± 645 |
| FM scores | 41 ± 16 |

**Figure 2:**
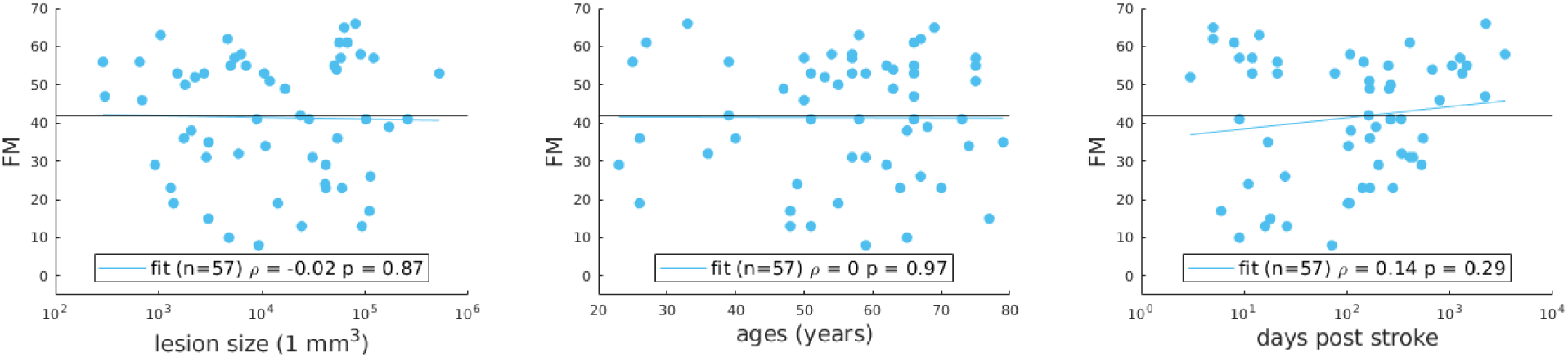
Correlation between FM scores and A. the lesion volume; B. age; and C. days post-stroke. The black line indicates FM score of 42, used to separate the group into H and L. The blue line is a linear fit for the variables.

Of the 57 patients, there are 8 patients on post-stroke days between 1 and 10 (L: 3; H: 5), 13 patients on post-stroke days between 10 and 100 (L: 7; H: 6), and 36 patients on post-stroke days beyond 100 (L: 18; H: 18).

### Structural connectivity

#### Structural disconnections involving the brainstem (n=57)

Figure 3 A shows the probability of each motor-related ROI being structurally disconnected from the brainstem when the patient is in the low FM score group. In the Figure, only the ROIs of the motor-related areas that are disconnected in at least one patient are shown. If a patient is in the LL group, we find the highest probability that ROIs in ipsilesional M1 are disconnected from the brainstem. However, disconnection of the brainstem from ipsilesional M1 is not itself an indicator of high probability of belonging to the LL group. The probability of low motor status when the ROI is disconnected from the brainstem is shown in Figure 3 B. Disconnection of the ROI from ipsilesional PMd to the brainstem is strongly predictive of being in the low FM score group. However, 23/24 patients having brainstem disconnected from at least one ROI within ipsilesional PMd also had a disconnection from at least one ROI in ipsilateral M1, indicating that this prediction reflects the joint disconnection of the brainstem from M1 and PMd. In addition, we observed a similarly high likelihood of belonging to the L group when brainstem disconnections involved M1 or PMd in either hemisphere.

**Figure 3:**
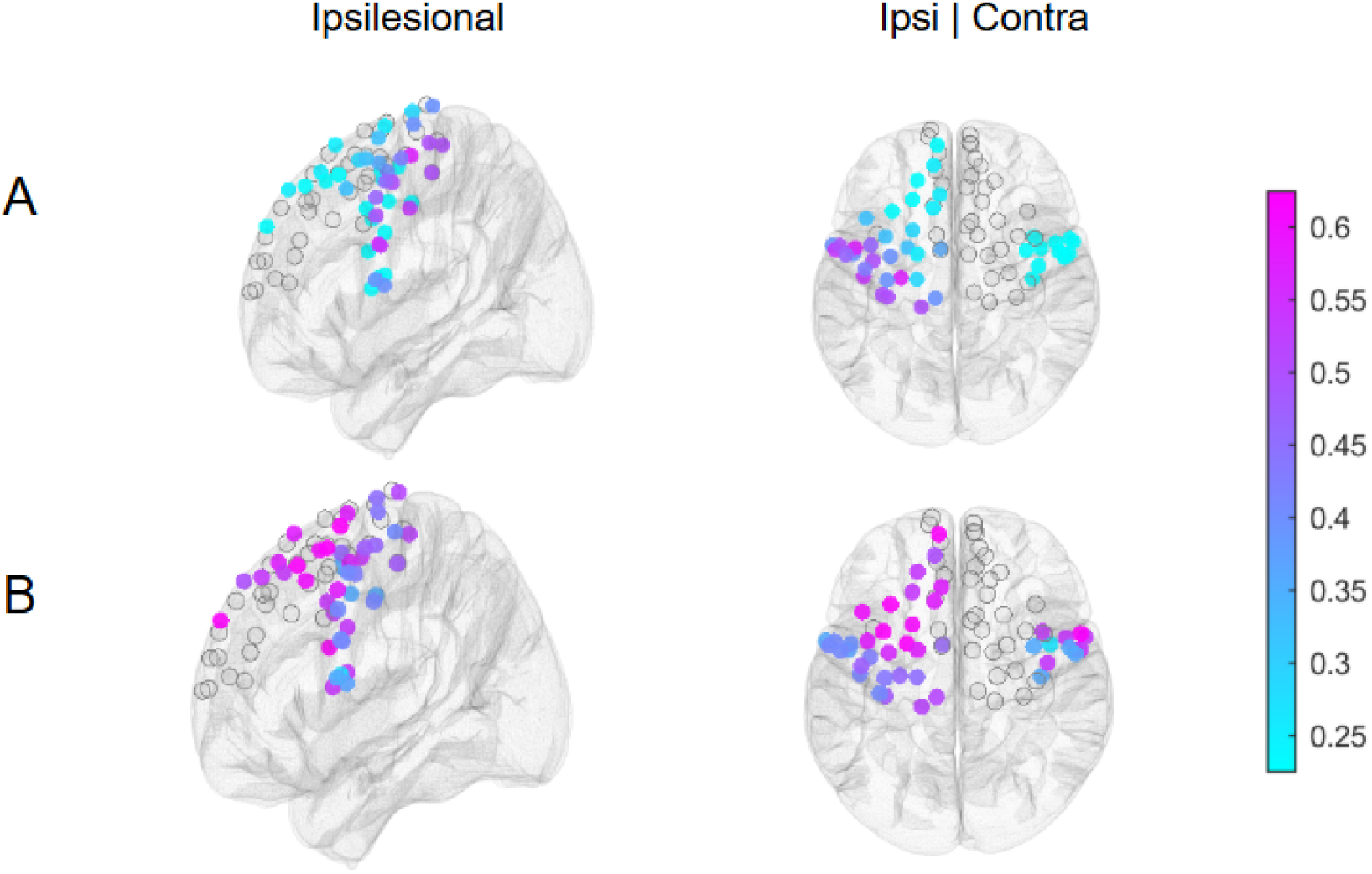
Structural Disconnections involving the Brainstem: A. The probability of the ROI being disconnected from the brainstem when the patient in in the low FM score (“L”) group *P*(*D*|*L*). The ROI in ipsilesional M1 has the highest probability. B. The probability of being in the low FM score group when the ROI is disconnected from the brainstem *P*(*L*|*D*). ROIs in the ipsilesional PMd have the highest probability.

#### Structural disconnections involving the motor-related ROIs (n=50)

We computed 3 network properties for motor-related ROIs (degree centrality, average efficiency, and betweenness centrality) from each patient’s reduced structural connectome. We split the patients into 2 groups according to their motor status – H vs L. Then we computed the ratio of their mean network measurement values in H/L across the 50 patients. We did a permutation test of H > L with a significance level of 0.05 to determine if disconnection of an ROI has significant influence on motor status. Motor-related ROIs with significant differences in these network properties are shown in Figure 4: The strongest effects (H > L) are found in the number of intact connections (degree centrality) of ipsilesional motor areas. In addition, the efficiency of communication of ipsilesional motor areas are all reduced in the L groups. The routing of signals through the motor areas (betweenness centrality) are more strongly influenced in M1 and PMd ROIs. For the contralesional motor cortex ROIs, degree centrality and betweenness centrality are reduced in the M1 and PMd ROIs in the L group.

**Figure 4:**
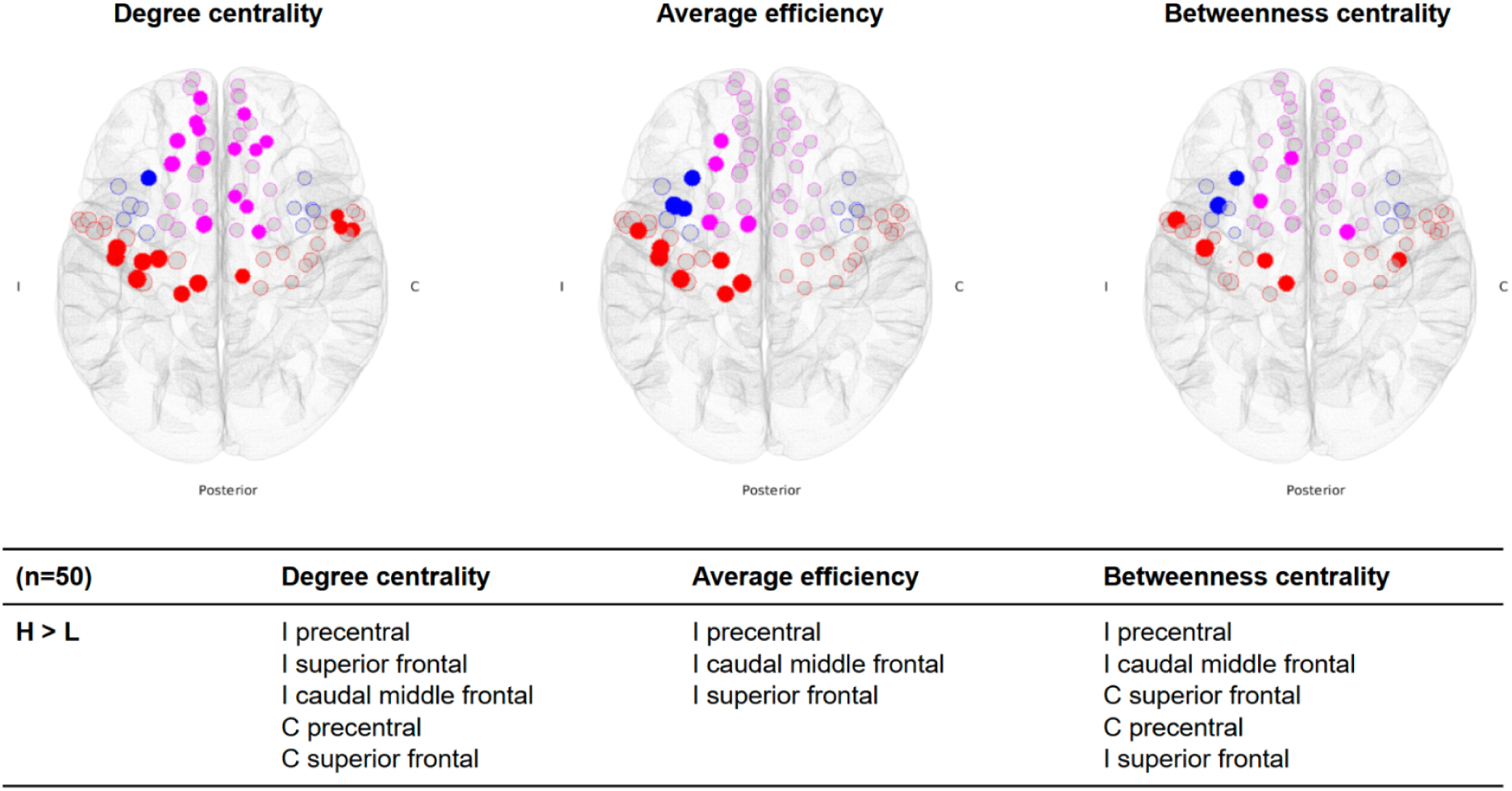
Structural network measurements in the motor-related areas. Magenta circle - SFG; Blue circle - PMd; Red circle – M1; I - ipsilesional hemisphere; C - contralesional hemisphere; H - high motor group; L - low motor group. Each color-filled solid circle represents a significant ROI with p < 0.05. Transparent circles are the remaining ROIs in each area. The ratios of the number of significant ROIs versus all ROIs in the motor-related areas are 27/78, 15/78, and 10/78 for degree centrality, average efficiency, and betweenness centrality, respectively. Areas with significant ROIs are listed in the table, in descending order of the ratio of each metric in the H and L group..

#### Structural disconnections involving ROIs outside motor areas (n=50)

We computed the motor betweenness centrality (see Method - Characterizing structural and functional networks using graph theory) for all ROIs outside of the motor-related areas from each patient’s reduced structural connectome. Due to stroke lesions, some ROIs may change in their centrality for the routing of signals from the motor areas. We did a two-tailed test with an alpha of 0.05 to determine if an ROI is significant, for which motor betweenness centrality is either higher in the H group (red dots) or L group (blue dots), as shown in Figure 5: Almost all ROIs that show an increase in motor betweenness centrality are found in the L group, indicating that new shortest pathways for the motor system to communicate with other cortical areas are associated with reduced FM scores. In simpler terms, patients with worse motor status tend to reroute signals through non-motor brain areas that are less efficient, suggesting a compensatory but inefficient network reorganization.

**Figure 5:**
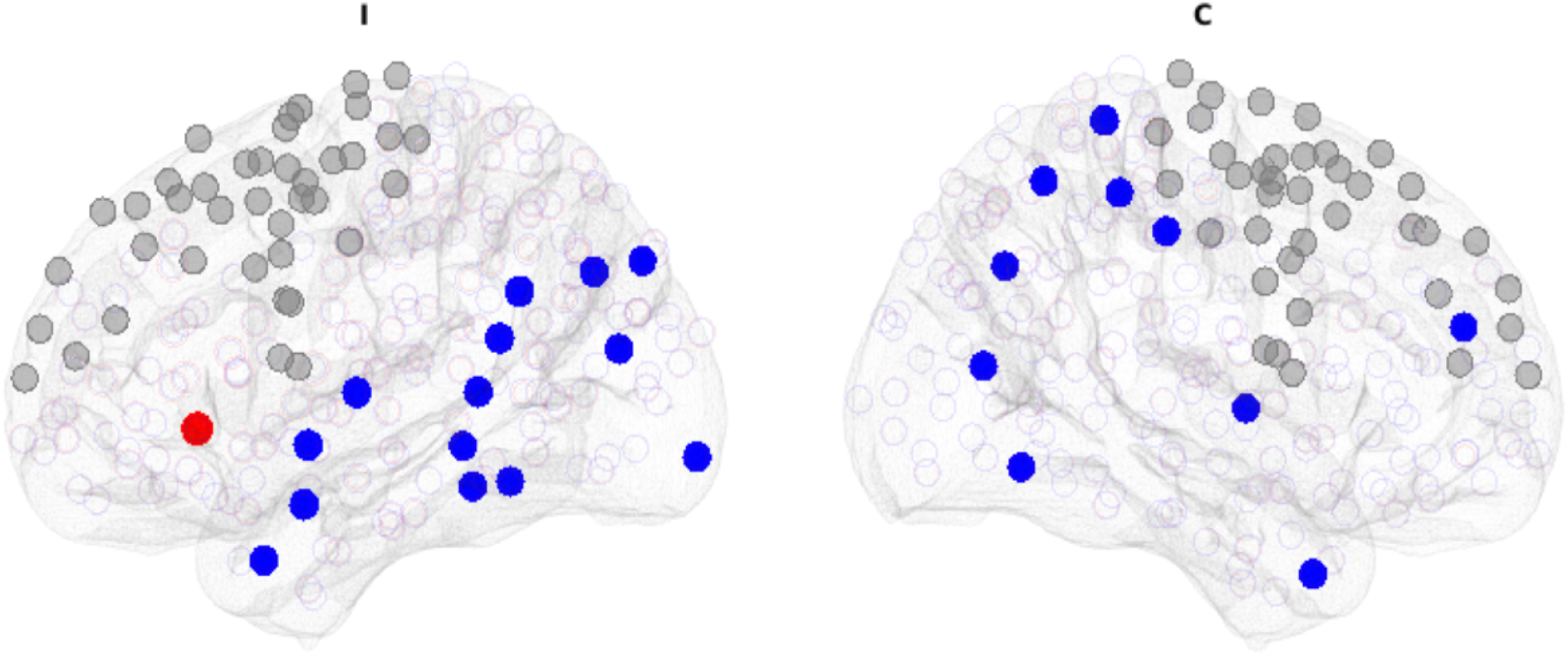
Significant ROIs outside the motor-related areas: Each color-filled solid circle represents a significant ROI (two-tailed, p 0.05). Transparent circles are non-significant ROIs. Sizes of circles correspond to the value of H/L or L/H. Red circle - H>L; Blue circle - L>H; Grey circle - Motor-related ROIs; I - ipsilesional hemisphere; C - contralesional hemisphere. The number of significant ROIs versus all connected ROIs outside of the motor-related areas are: 1/370 for H>L; 24/370 for L>H. Only ipsilesional lateral orbitofrontal showed significant H>L. Areas of significant ROIs with L>H in order of effect size: contralesional pallidum, fusiform, inferior parietal lobule, lateral occipital cortex, middle temporal gyrus, paracentral lobule, precuneus, rostral middle frontal gyrus, supramarginal gyrus, and ipsilesional banks of the superior temporal sulcus, fusiform gyrus, inferior parietal lobule, inferior temporal gyrus, lateral occipital cortex, middle temporal gyrus, parahippocampal gyrus, superior parietal lobule, and superior temporal gyrus.

### EEG functional connectivity

#### EEG functional connectivity involving motor-related ROIs

We analyzed resting state EEG data from all 57 patients, source-localized and aggregated them into 448 ROIs according to the Lausanne parcellation. Using the AGL informed by each patient’s reduced structure connectome and the cross-validation procedure, we estimated the partial coherence in each of 6 EEG frequency bands in order to compute 3 network properties (degree centrality, average efficiency, and betweenness centrality) for all 78 ROIs in the ipsilesional and contralesional motor areas (M1, PMd, and SFG). The motor-related areas with significant ROIs for the δ, α, β_1_, and β_2_ frequency bands are shown in Table [tab:E3TB] with δ, β_1_ and β_2_ displayed in Figure 6. All of the network measures were elevated in the H group compared to the L group, in all 78 ROIs, indicating a globally strengthened motor network organization associated with better motor function. The main significant effect was elevated network function in the H group in the δ band, where each network measure was higher in every ROI, with the strongest effects found in ipsilesional M1. All three ipsilesional motor areas showed significantly elevated degree centrality and efficiency in the H group, indicating that ipsilesional M1 forms a greater number of functional connections across the brain in the delta band, and that these connections operate with increased efficiency. Similarly elevated network measures were found in the contralesional SFG in the H group. Elevated network measures were also found in every ROI in the β band in the H group, with significant elevation of all 3 measures in the contralesional M1 and SFG. In the α band, elevated betweenness centrality was observed in all 3 contralesional motor areas in the H group.

**Figure 6:**
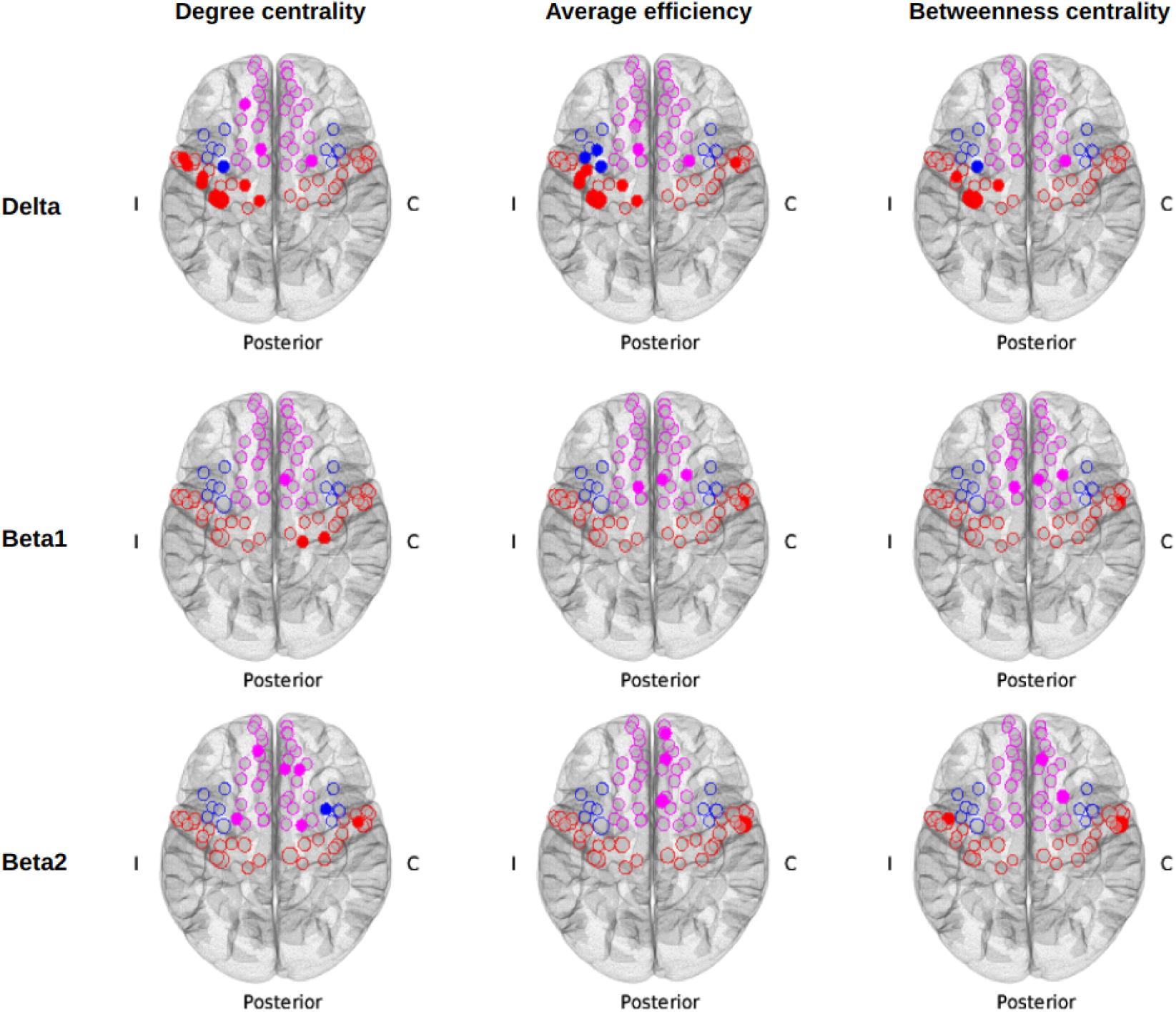
Significant ROIs in the motor-related areas for δ and β frequencies: All color-filled dots and transparent circles are the ROIs with network metrics such that H>L. Each color-filled dot represents a significant ROI with p < 0.05 (two-tailed). There was no ROI with L>H for any network measures. Sizes of dots and circles correspond to the value of H/L. Magenta - superior frontal; Blue - caudal middle frontal; Red - precentral; I - ipsilesional hemisphere; C - contralesional hemisphere.

#### EEG functional connectivity involving ROIs outside motor areas (n=57)

Brain areas with significant ROIs in motor betweenness centrality were found primarily in the δ (12 ROIs), β_1_ (11 ROIs), and β_2_ (17 ROIs) frequency bands and are listed in Table 2 and displayed in Figure 7. We predominantly observed higher motor betweenness in the LL group in the δ and β_2_ bands and higher motor betweenness in the H group in the β_1_ band. Based on the spatial location of the significant ROIs for H*>*L and L*>*H, higher FM scores are likely associated with higher motor betweenness centrality in areas near the ipsilesional motor areas. Whereas lower FM scores are likely associated with higher motor betweenness centrality in brain regions further away from the motor related areas. In sum, better motor status is associated with more efficient routing through nearby motor-related areas, while worse motor status involves detouring through more distant and less optimal regions.

**Figure 7:**
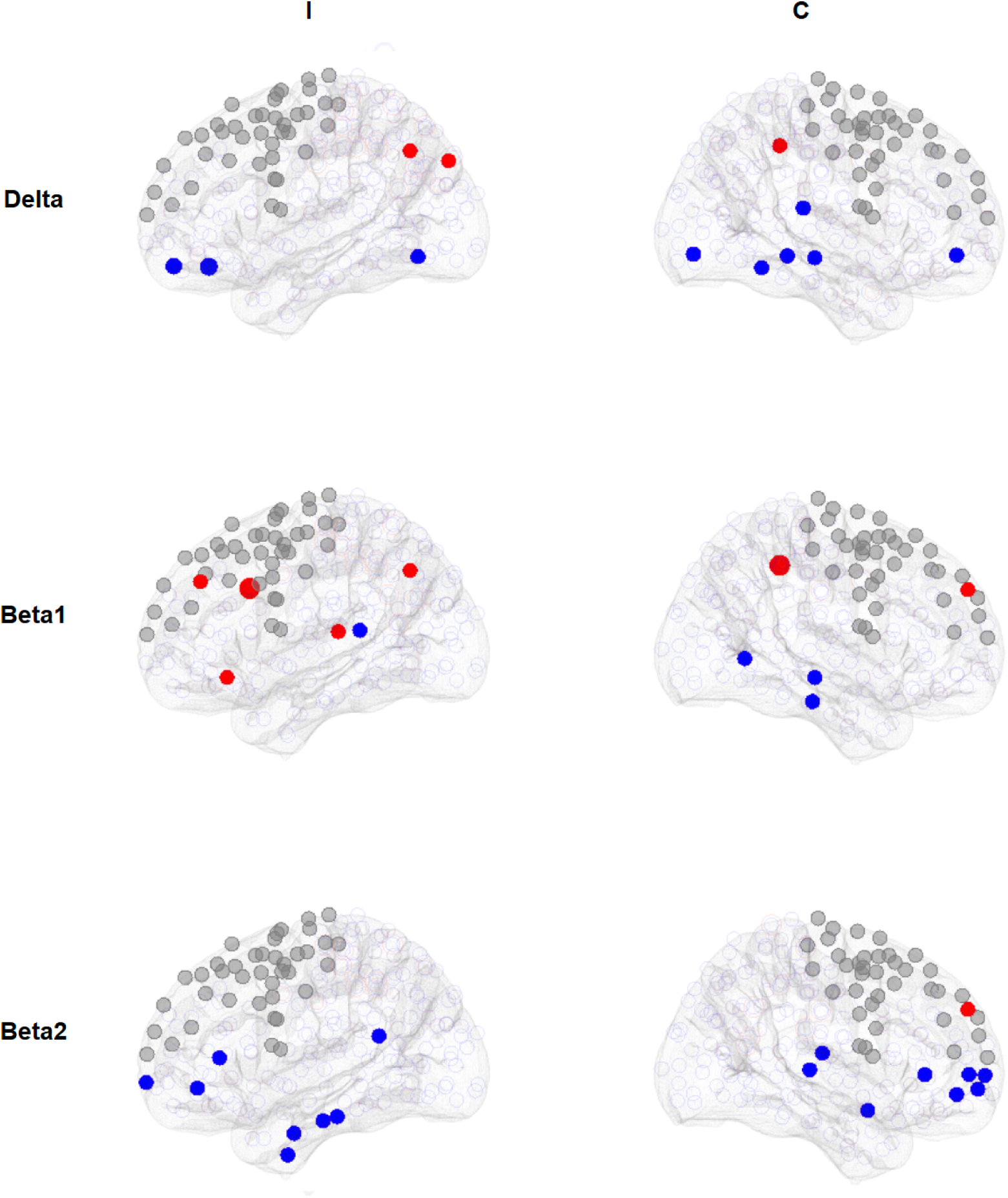
Significant ROIs outside the motor-related areas in δ and β frequencies (Left and Right view of the ipsilesional and contralesional hemisphere): Each color-filled dot represents a significant ROI with p 0.05 (two-tailed). Open-colored circles are non-significant ROIs. The sizes of dots and circles correspond to the values of H/L or L/H. Red - H/L 1; Blue - L/H 1; I - ipsilesional hemisphere; C - contralesional hemisphere.

**Table 2:** Brain areas of significant ROIs outside of the motor-related areas in motor betweenness centrality found in δ, β1, and β2 bands: I - ipsilesional hemisphere; C - contralesional hemisphere. Areas with significant ROIs are listed in descending order of effect size. ROIs were considered significant with p < 0.05 (two-tailed). The number of significant ROIs versus all connected ROIs outside of the motor-related areas for H > L and L > H respectively, are: 3/370 and 9/370 in δ, 7/370 and 4/370 in β1, 1/370 and 16/370 in β2.

|  | Delta | Beta1 | Beta2 |
| --- | --- | --- | --- |
| **H > L** | C precuneus<br>I inferior parietal | C precuneus<br>I caudal anterior cingulate<br>C rostral middle frontal<br>I lateral orbitofrontal<br>I rostral middle frontal<br>I inferior parietal<br>I superior temporal | C rostral middle frontal |
| **L > H** | I lateral orbitofrontal<br>C lingual<br>C superior temporal<br>C medial orbitofrontal<br>C parahippocampal<br>I fusiform<br>C middle temporal<br>C fusiform | C inferior temporal<br>C middle temporal<br>I bankssts | C medial orbitofrontal<br>C pars triangularis<br>C rostral middle frontal<br>C superior temporal<br>C transverse temporal<br>I lateral orbitofrontal<br>I frontal pole<br>I pars triangularis<br>I fusiform<br>I parahippocampal<br>I entorhinal<br>I inferior temporal<br>I bankssts |

## Discussion

### Structural Connectivity Findings

Our study revealed that structural disconnection patterns, particularly involving motor-related areas, robustly distinguished motor status post-stroke. Structural disconnection between the brainstem and ipsilesional PMd and M1 was the strongest predictor of poor motor status. This is not too surprising as it likely reflects disruption of descending motor pathways (such as the corticospinal tract) as well as ascending sensory tracts, both critical to motor function. In addition, lower degree centrality, average efficiency, and betweenness centrality in ipsilesional motor areas (M1, PMd, and SFG) were consistently associated with lower motor status. This pattern suggests that reduced degree centrality indicates fewer direct connections between ipsilesional motor regions and the rest of the network, lower efficiency reflects less effective communication across distributed brain areas, and diminished betweenness centrality signifies a reduced hub-like role in routing information. Together, these changes imply a loss of integrative capacity and weakened influence of motor regions within the broader network, limiting their ability to coordinate distributed activity necessary for motor function. These findings align with previous studies highlighting the importance of both structural and functional integrity of the motor system in determining functional outcomes after stroke (Olafson et al. 2021; L. Wang et al. 2010; Wanni Arachchige et al. 2021).

Outside of motor areas, we observed that patients with lower motor status had increased motor betweenness centrality in distant cortical regions. This indicates that after significant motor system disruption, communication reroutes through more distant brain areas, potentially reflecting maladaptive plasticity, as previously suggested in models of stroke recovery(Takeuchi and Izumi 2012; Alia et al. 2017). Thus, preservation of direct motor pathways supports better motor status, while reliance on alternative, less efficient routes correlates with worse motor outcomes.

### EEG Functional Connectivity Findings

EEG-based functional connectivity provided complementary insights. In the δ band (1-3 Hz), patients with better motor status exhibited higher degree centrality, efficiency, and betweenness centrality in ipsilesional motor regions, indicating indicating greater local connectivity, more efficient information transfer, and stronger hub-like roles within the recovering motor network. This mirrors the structural findings and supports prior work showing the relevance of low-frequency oscillations in capturing functional recovery dynamics(Cassidy et al. 2020).

In higher frequencies, particularly the β band (14-29 Hz), enhanced functional connectivity was seen in contralesional motor areas in patients with better outcomes. This finding is consistent with evidence that contralesional motor areas can compensate for ipsilesional damage in stroke recovery(Rehme et al. 2012; Buetefisch 2015 and Cramer et al., 1997). The utility of increased contralesional hemisphere activity to successful recovery varies according to severity of the stroke. Current results suggest that, among the broad finding of increased contralesional activity, favorable contributions can be identified using the β band functional connectivity.

Outside motor regions, motor betweenness centrality patterns in EEG showed a spatial shift: in the β_1_ band (14-20 Hz), greater routing through regions near ipsilesional M1 correlated with better motor status, while greater routing through distant cortical areas (in δ and β_2_ bands) correlated with worse outcomes. These patterns reinforce that efficient local reorganization, rather than global network rerouting, supports better recovery, here in the motor system.

### Comparison of Areas Identified in EEG and Structural Connectome

There was notable overlap between the structural and functional connectivity findings. In particular, ipsilesional motor areas, especially M1 and PMd, were consistently identified as critical hubs in both modalities. Structural disconnection of these areas predicted poor motor status, while higher EEG connectivity in these regions in δ and β bands predicted better outcomes. This concordance suggests that EEG-based metrics, particularly δ band functional connectivity, reflect the underlying structural integrity and reorganization of motor networks post-stroke. Differences also emerged: while structural disconnections primarily indicated the extent of injury, EEG revealed plastic changes over time, highlighting new hubs for motor signal propagation that were not visible structurally (Burke et al., 2015; Nouri and Cramer, 2011; Wu et al., 2015; and Ingemanson et al., 2019). This emphasizes the complementary value of combining structural and EEG analyses (Babaeeghazvini et al. 2021).

### Neural plasticity in motor recovery after stroke

The recovery process after a stroke involves the growth of axons and dendrites, formation of new synapses, and the reweighting of alternative neuronal circuits or brain networks (Wieloch and Nikolich 2006; Alhadidi et al. 2024). Additionally, studies of human biopsies and autopsies indicate that neuronal migration occurs after a stroke (Jin et al. 2006), though the clinical relevance of this in humans remains uncertain.

Consequently, it is reasonable to suggest that the neural network changes during post-stroke motor recovery, not only within but also outside the motor-related areas, reflect clinically relevant neural plasticity. The motor betweenness centrality measurement we designed in this study is well-suited for examining such plasticity occurring outside the motor areas in facilitating motor brain-wide communication related to motor system function after stroke.

### Implication of motor network characteristics

The structural results show that degree centrality identifies the largest number of regions of interest, underscoring how important the overall number of connections is for motor status. Both degree centrality and betweenness centrality reflect the influence of the contralesional motor network: higher values mean that these regions are better positioned to interact with and shape activity across the network, which supports better motor outcomes. By contrast, average efficiency is more sensitive to localized changes on the ipsilesional side. When more direct connections between ipsilesional motor areas are preserved, efficiency increases, allowing information to travel more effectively within this local network, and this is linked to better motor recovery.

### Connectivity drives stroke recovery

This study satisfies Bradford Hill’s nine tenets in supporting the plausibility of a causal relationship between brain connectivity and motor recovery. Bradford Hill’s nine tenets (Hill 1965), often referred to as “Hill’s criteria of causation,” are a group of principles that provide a systematic approach to evaluating the plausibility of a causal relationship in connectivity research in stroke (Cassidy, Mark, and Cramer 2022). The support for the nine tenets are: (i) Strength - Larger associations of ROIs with higher probability or smaller p values are found with the motor status; (ii) Consistency - Data include patients across different stroke stages from acute and subacute to chronic status; also, neuroimaging modalities derived from both structural MRI and functional EEG are convergent; (iii) Specificity - Distinct associations between lesion-related damage to ipsilesional motor areas and recovery status; (iv) Temporality - Changes in structural connectivity follow stroke precede the functional connectivity from EEG and the motor recovery status; (v) Biological gradient - Magnitude of connectivity change was related to severity of motor deficit (H/L or L/H); (vi) Plausibility and (vii) Coherence - The ipsilesional motor areas with their nearby regions and compensatory enhancement of their contralesional counterparts being most important to motor recovery are sensible and concordant when viewed in the context of the current knowledge of brain physiology and anatomy after stroke; (viii) Experiment - Changes defined here are related to stroke, in comparison to healthy controls, and so reflect two different disease states, albeit not experimentally assigned; (ix) Analogy - Similar associations of connectivity changes in the ipsilesional (Rehme et al. 2011; Grefkes et al. 2008; Ingemanson et al. 2019) and contralesional motor areas (Liu et al. 2015; Cassidy et al. 2020) have been reported in relation to motor recovery after stroke. Collectively, the overall weight of the evidence from these 9 tenets supports our interpretation of the causal relationship between connectivity and motor status.

### Meaning and Clinical Relevance

Together, structural disconnections delineate the initial severity of network disruption, while EEG connectivity captures the subsequent functional adaptations. Structural measures, such as reduced degree and efficiency in ipsilesional motor areas, reflect stroke-related injury and so may be able to identify patients at higher risk for poor recovery based on studies performed acutely, and so could potentially identify patients in whom standard dose rehabilitation interventions are less likely to be efficacious. EEG measures provide dynamic biomarkers of functional plasticity that can track progress and predict rehabilitation response(Cassidy et al. 2020; Keser et al. 2022; Wu et al., 2015; Zhou et al., 2018) and so may be useful to monitor functional connectivity. Such a dynamic biomarker for post-stroke rehabilitation therapy would be analogous to what is done currently in many branches of medicine, e.g., serial measurement of serum TSH levels to guide levothyroxine treatment of hypothyroidism or serial measurement of the cell population of interest to treat various myeloproliferative disorders.

Clinically, these findings suggest that therapies aimed at strengthening ipsilesional motor networks and facilitating compensatory contralesional activation, especially targeting δ and β band oscillations, may enhance recovery(Cassidy et al. 2020; Espenhahn et al. 2020). Additionally, avoiding maladaptive recruitment of distant, non-motor regions could be a therapeutic goal.

### Limitations

Several limitations exist in this work. There are edges that we failed to account for because our structural connectome did not include some genuine anatomical connections, such as those to the cerebellum, which is crucial for motor coordination. After aggregating the source-localized EEG, we removed the subcortical ROIs, as we believe that scalp EEG is more reflective of electrical activities at the cortical level.

However, the exclusion of subcortical activity means we missed important components of neural activity that mediate motor coordination in subcortical brain structures such as the thalamus and basal ganglia.

In our coherence measurement, both phase and amplitude were considered when computing functional connectivity. Other studies suggest that there may be potentially separate mechanisms at play in generating neuronal coupling of phase and amplitude (Siems and Siegel 2018; Engel et al. 2013; Cabral et al. 2014).

### Summary

We implemented several methodological refinements to improve EEG-based network analysis in stroke. First, to overcome limitations in prior source models (e.g., Wodeyar & Srinivasan, 2022), we developed a high-resolution forward model in MNE using surface-constrained dipoles with fixed orientations, optimized for our 256-channel EEG data. For estimating source activity, we tested multiple inverse solutions and selected a weighted L2 norm method based on its superior reconstruction of scalp EEG signals. To estimate effective connectivity, we used a cross-validated complex-valued Gaussian graphical model (cGGM) informed by each participant’s individualized binarized structural connectome. This approach allowed us to estimate partial coherence—capturing conditional dependencies in both amplitude and phase—offering greater stability and specificity than partial correlation (Colclough et al., 2016).

We find that structural connectivity damage within motor circuits defines the starting point for motor impairment, while EEG functional connectivity reflects dynamics of motor status and its recovery over time or in response to treatment. Measuring both provides a comprehensive, actionable framework for prognosis and rehabilitation therapy targeting after stroke.

In summary, the structural and EEG functional network properties of degree centrality, average efficiency, and betweenness centrality are reliable methods for identifying significant ROIs in motor-related areas that predict and indicate motor status in recovery from stroke. In contrast, motor betweenness reflects the contribution of the alternative paths and neuroplasticity outside of motor-related areas. The δ and β frequencies better characterize EEG networks related to motor status.

## Data Availability

All data produced in the present study are available upon reasonable request to the authors

## Notes

### Competing Interest Statement

Dr. Cramer serves as a consultant for Astellas, Bayer, Beren Therapeutics, BlueRock Therapeutics, BrainQ, Constant Therapeutics, Medtronic, MicroTransponder, Myomo, NeuroTrauma Sciences, Simcere, and TRCare. Other authors declare no conflict of interest.

### Funding Statement

This study was funded by NSF Award #: 2126976

### Author Declarations

This study protocol received approvals from the University of California, Irvine, and the University of California, Los Angeles Institutional Review Board.

## Reference

Alhadidi, Qasim M, Ghaith A Bahader, Oiva Arvola, Philip Kitchen, Zahoor A Shah, and Mootaz M Salman. 2024. “Astrocytes in Functional Recovery Following Central Nervous System Injuries.” The Journal of Physiology 602 (13): 3069–96.

Alia, Claudia, Cristina Spalletti, Stefano Lai, Alessandro Panarese, Giuseppe Lamola, Federica Bertolucci, Fabio Vallone, et al. 2017. “Neuroplastic Changes Following Brain Ischemia and Their Contribution to Stroke Recovery: Novel Approaches in Neurorehabilitation.” Frontiers in Cellular Neuroscience 11: 76.

Babaeeghazvini, Parinaz, Laura M Rueda-Delgado, Jolien Gooijers, Stephan P Swinnen, and Andreas Daffertshofer. 2021. “Brain Structural and Functional Connectivity: A Review of Combined Works of Diffusion Magnetic Resonance Imaging and Electro-Encephalography.” Frontiers in Human Neuroscience 15: 721206.

Buetefisch, Cathrin M. 2015. “Role of the Contralesional Hemisphere in Post-Stroke Recovery of Upper Extremity Motor Function.” Frontiers in Neurology 6: 214.

Bullmore, Ed, and Olaf Sporns. 2012. “The Economy of Brain Network Organization.” Nature Reviews Neuroscience 13 (5): 336.

Burke, Erin, Lucy Dodakian, Jill See, Alison McKenzie, Jeff D Riley, Vu Le, and Steven C Cramer. 2014. “A Multimodal Approach to Understanding Motor Impairment and Disability After Stroke.” Journal of Neurology 261: 1178–86.

Burke Quinlan, E., Dodakian, L., See, J., McKenzie, A., Le, V., Wojnowicz, M., Shahbaba, B., and Cramer, S. C. (2015). “Neural function, injury, and stroke subtype predict treatmentc gains after stroke.” Annals of neurology, 77(1):132–145.

Cabral, Joana, Henry Luckhoo, Mark Woolrich, Morten Joensson, Hamid Mohseni, Adam Baker, Morten L Kringelbach, and Gustavo Deco. 2014. “Exploring Mechanisms of Spontaneous Functional Connectivity in MEG: How Delayed Network Interactions Lead to Structured Amplitude Envelopes of Band-Pass Filtered Oscillations.” Neuroimage 90: 423–35.

Cammoun, Leila, Xavier Gigandet, Djalel Meskaldji, Jean Philippe Thiran, Olaf Sporns, Kim Q Do, Philippe Maeder, Reto Meuli, and Patric Hagmann. 2012. “Mapping the Human Connectome at Multiple Scales with Diffusion Spectrum MRI.” Journal of Neuroscience Methods 203 (2): 386–97.

Campos, Baruc, Hoseok Choi, Andrew T DeMarco, Anna Seydell-Greenwald, Sara J Hussain, Mary T Joy, Peter E Turkeltaub, and William Zeiger. 2023. “Rethinking Remapping: Circuit Mechanisms of Recovery After Stroke.” Journal of Neuroscience 43 (45): 7489–7500.

Cassidy, Jessica M, Jasper I Mark, and Steven C Cramer. 2022. “Functional Connectivity Drives Stroke Recovery: Shifting the Paradigm from Correlation to Causation.” Brain 145 (4): 1211–28.

Cassidy, Jessica M, George Tran, Erin B Quinlan, and Steven C Cramer. 2018. “Neuroimaging Identifies Patients Most Likely to Respond to a Restorative Stroke Therapy.” Stroke 49 (2): 433–38.

Cassidy, Jessica M, Anirudh Wodeyar, Ramesh Srinivasan, and Steven C Cramer. 2021. “Coherent Neural Oscillations Inform Early Stroke Motor Recovery.” Human Brain Mapping 42 (17): 5636–47.

Cassidy, Jessica M, Anirudh Wodeyar, Jennifer Wu, Kiranjot Kaur, Ashley K Masuda, Ramesh Srinivasan, and Steven C Cramer. 2020. “Low-Frequency Oscillations Are a Biomarker of Injury and Recovery After Stroke.” Stroke 51 (5): 1442–50.

Colby, John B, Lindsay Soderberg, Catherine Lebel, Ivo D Dinov, Paul M Thompson, and Elizabeth R Sowell. 2012. “Along-Tract Statistics Allow for Enhanced Tractography Analysis.” Neuroimage 59 (4): 3227–42.

Colclough, Giles L, Mark W Woolrich, PK Tewarie, Matthew J Brookes, Andrew J Quinn, and Stephen M Smith. 2016. “How Reliable Are MEG Resting-State Connectivity Metrics?” Neuroimage 138: 284–93.

Cramer, Steven C., Gereon Nelles, Randall R. Benson, Jill D. Kaplan, Robert A. Parker, Ken K. Kwong, David N. Kennedy, Seth P. Finklestein, and Bruce R. Rosen. “A functional MRI study of subjects recovered from hemiparetic stroke.” Stroke 28, no. 12 (1997): 2518–2527.

Daducci, Alessandro, Stephan Gerhard, Alessandra Griffa, Alia Lemkaddem, Leila Cammoun, Xavier Gigandet, Reto Meuli, Patric Hagmann, and Jean-Philippe Thiran. 2012. “The Connectome Mapper: An Open-Source Processing Pipeline to Map Connectomes with MRI.” PloS One 7 (12): e48121.

Engel, Andreas K, Christian Gerloff, Claus C Hilgetag, and Guido Nolte. 2013. “Intrinsic Coupling Modes: Multiscale Interactions in Ongoing Brain Activity.” Neuron 80 (4): 867–86.

Espenhahn Svenja, Holly E Rossiter, Bernadette CM van Wijk, Nell Redman, Jane M Rondina, Joern Diedrichsen, and Nick S Ward. 2020. “Sensorimotor Cortex Beta Oscillations Reflect Motor Skill Learning Ability After Stroke.” Brain Communications 2 (2): fcaa161.

Fischl, Bruce. 2012. “FreeSurfer.” Neuroimage 62 (2): 774–81.

Fornito, Alex, Andrew Zalesky, and Edward Bullmore. 2016. Fundamentals of Brain Network Analysis. Academic Press.

Friedman, Jerome, Trevor Hastie, and Robert Tibshirani. 2008. “Sparse Inverse Covariance Estimation with the Graphical Lasso.” Biostatistics 9 (3): 432–41.

Fries, Pascal. 2005. “A Mechanism for Cognitive Dynamics: Neuronal Communication Through Neuronal Coherence.” Trends in Cognitive Sciences 9 (10): 474–80.

Gladstone, David J, Cynthia J Danells, and Sandra E Black. 2002. “The Fugl-Meyer Assessment of Motor Recovery After Stroke: A Critical Review of Its Measurement Properties.” Neurorehabilitation and Neural Repair 16 (3): 232–40.

Goñi Joaquın, Martijn P van den Heuvel, Andrea Avena-Koenigsberger, Nieves Velez de Mendizabal, Richard F Betzel, Alessandra Griffa, Patric Hagmann, Bernat Corominas-Murtra, Jean-Philippe Thiran, and Olaf Sporns. 2014. “Resting-Brain Functional Connectivity Predicted by Analytic Measures of Network Communication.” Proceedings of the National Academy of Sciences 111 (2): 833–38.

Gramfort, Alexandre, Martin Luessi, Eric Larson, Denis A. Engemann, Daniel Strohmeier, Christian Brodbeck, Roman Goj, et al. 2013. “MEG and EEG Data Analysis with MNE-Python.” Frontiers in Neuroscience 7 (267): 1–13. 10.3389/fnins.2013.00267.

Grefkes, Christian, Dennis A Nowak, Simon B Eickhoff, Manuel Dafotakis, Jutta Küst, Hans Karbe, and Gereon R Fink. 2008. “Cortical Connectivity After Subcortical Stroke Assessed with Functional Magnetic Resonance Imaging.” Annals of Neurology 63 (2): 236–46.

Griffis, J. C., Metcalf, N. V., Corbetta, M., & Shulman, G. L. (2019). Structural disconnections explain brain network dysfunction after stroke. Cell reports, 28(10), 2527–2540.

Hagberg, Aric A., Daniel A. Schult, and Pieter J. Swart. 2008. “Exploring Network Structure, Dynamics, and Function Using NetworkX.” In Proceedings of the 7th Python in Science Conference, edited by Gaël Varoquaux, Travis Vaught, and Jarrod Millman, 11–15. Pasadena, CA USA.

Hill, Austin Bradford. 1965. “The Environment and Disease: Association or Causation?” Sage Publications.

Hsieh, Cho-Jui, Inderjit S Dhillon, Pradeep K Ravikumar, and Mátyás A Sustik. 2011. “Sparse Inverse Covariance Matrix Estimation Using Quadratic Approximation.” In Advances in Neural Information Processing Systems, 2330–38.

Imayoshi, Itaru, Masayuki Sakamoto, Toshiyuki Ohtsuka, Keizo Takao, Tsuyoshi Miyakawa, Masahiro Yamaguchi, Kensaku Mori, Toshio Ikeda, Shigeyoshi Itohara, and Ryoichiro Kageyama. 2008. “Roles of Continuous Neurogenesis in the Structural and Functional Integrity of the Adult Forebrain.” Nature Neuroscience 11 (10): 1153–61.

Ingemanson, Morgan L, Justin R Rowe, Vicky Chan, Eric T Wolbrecht, David J Reinkensmeyer, and Steven C Cramer. 2019. “Somatosensory System Integrity Explains Differences in Treatment Response After Stroke.” Neurology 92 (10): e1098–108.

Ismail, Umi Nabilah, Noorazrul Yahya, and Hanani Abdul Manan. 2024. “Investigating Functional Connectivity Related to Stroke Recovery: A Systematic Review.” Brain Research 1840: 149023.

Jenkinson, Mark, Christian F Beckmann, Timothy EJ Behrens, Mark W Woolrich, and Stephen M Smith. 2012. “Fsl.” Neuroimage 62 (2): 782–90.

Jin, Kunlin, Xiaomei Wang, Lin Xie, Xiao Ou Mao, Wei Zhu, Yin Wang, Jianfeng Shen, Ying Mao, Surita Banwait, and David A Greenberg. 2006. “Evidence for Stroke-Induced Neurogenesis in the Human Brain.” Proceedings of the National Academy of Sciences 103 (35): 13198–202.

Kantak, S. S., Stinear, J. W., Buch, E. R., and Cohen, L. G. (2012). “Rewiring the brain: potential role of the premotor cortex in motor control, learning, and recovery of function following brain injury”. Neurorehabilitation and neural repair, 26(3):282–292.

Keser, Zafer, Samuel C Buchl, Nathan A Seven, Matej Markota, Heather M Clark, David T Jones, Giuseppe Lanzino, Robert D Brown Jr, Gregory A Worrell, and Brian N Lundstrom. 2022. “Electroencephalogram (EEG) with or Without Transcranial Magnetic Stimulation (TMS) as Biomarkers for Post-Stroke Recovery: A Narrative Review.” Frontiers in Neurology 13: 827866.

Koch, Philipp J, Chang-Hyun Park, Gabriel Girard, Elena Beanato, Philip Egger, Giorgia Giulia Evangelista, Jungsoo Lee, et al. 2021. “The Structural Connectome and Motor Recovery After Stroke: Predicting Natural Recovery.” Brain 144 (7): 2107–19.

Koh, Chia-Lin, Chun-Hung Yeh, Xiaoyun Liang, Rishma Vidyasagar, Rüdiger J Seitz, Michael Nilsson, Alan Connelly, and Leeanne M Carey. 2021. “Structural Connectivity Remote from Lesions Correlates with Somatosensory Outcome Poststroke.” Stroke 52 (9): 2910–20.

Kuceyeski A., Maruta J., Relkin N., & Raj A. (2013). The network modification (nemo) tool: elucidating the effect of white matter integrity changes on cortical and subcortical structural connectivity. Brain Connectivity, 3(5), 451–463.

Leker, Ronen R, Frank Soldner, Ivan Velasco, Denise K Gavin, Andreas Androutsellis-Theotokis, and Ronald DG McKay. 2007. “Long-Lasting Regeneration After Ischemia in the Cerebral Cortex.” Stroke 38 (1): 153–61.

Lepage, K. Q., Kramer, M. A., & Eden, U. T. (2013). Some sampling properties of common phase estimators. Neural computation, 25(4), 901–921.

Lim, Jae-Sung, and Dong-Wha Kang. 2015. “Stroke Connectome and Its Implications for Cognitive and Behavioral Sequela of Stroke.” Journal of Stroke 17 (3): 256.

Lin, D. J., Cloutier, A. M., Erler, K. S., Cassidy, J. M., Snider, S. B., Ranford, J., Parlman, K., Giatsidis, F., Burke, J. F., Schwamm, L. H., et al. (2019). “Corticospinal tract injury estimated from acute stroke imaging predicts upper extremity motor recovery after stroke.” Stroke, 50(12):3569–3577.

Liu, Jingchun, Wen Qin, Jing Zhang, Xuejun Zhang, and Chunshui Yu. 2015. “Enhanced Interhemispheric Functional Connectivity Compensates for Anatomical Connection Damages in Subcortical Stroke.” Stroke 46 (4): 1045–51.

Meder, D. and Siebner, H. R. (2018). “Spectral signatures of neurodegenerative diseases: how to decipher them?” Brain, 141(8):2241–2244.

Mišić, Bratislav, Richard F Betzel, Azadeh Nematzadeh, Joaquin Goñi, Alessandra Griffa, Patric Hagmann, Alessandro Flammini, Yong-Yeol Ahn, and Olaf Sporns. 2015. “Cooperative and Competitive Spreading Dynamics on the Human Connectome.” Neuron 86 (6): 1518–29.

Nouri, S., & Cramer, S. C. (2011). Anatomy and physiology predict response to motor cortex stimulation after stroke. Neurology, 77(11), 1076–1083.

Nunez, Paul L, and Ramesh Srinivasan. 2006. Electric Fields of the Brain: The Neurophysics of EEG. Oxford University Press, USA.

Ohab, John J, Sheila Fleming, Armin Blesch, and S Thomas Carmichael. 2006. “A Neurovascular Niche for Neurogenesis After Stroke.” Journal of Neuroscience 26 (50): 13007–16.

Olafson, Emily R, Keith W Jamison, Elizabeth M Sweeney, Hesheng Liu, Danhong Wang, Joel E Bruss, Aaron D Boes, and Amy Kuceyeski. 2021. “Functional Connectome Reorganization Relates to Post-Stroke Motor Recovery and Structural and Functional Disconnection.” Neuroimage 245: 118642.

Paul, Theresa, Valerie M Wiemer, Lukas Hensel, Matthew Cieslak, Caroline Tscherpel, Christian Grefkes, Scott T Grafton, Gereon R Fink, and Lukas J Volz. 2023. “Interhemispheric Structural Connectivity Underlies Motor Recovery After Stroke.” Annals of Neurology 94 (4): 785–97.

Rathore, S. S., Hinn, A. R., Cooper, L. S., Tyroler, H. A., & Rosamond, W. D. (2002). Characterization of incident stroke signs and symptoms: findings from the atherosclerosis risk in communities study. Stroke, 33(11), 2718–2721.

Rehme, Anne K, Simon B Eickhoff, Claudia Rottschy, Gereon R Fink, and Christian Grefkes. 2012. “Activation Likelihood Estimation Meta-Analysis of Motor-Related Neural Activity After Stroke.” Neuroimage 59 (3): 2771–82.

Rehme, Anne K, Simon B Eickhoff, Ling E Wang, Gereon R Fink, and Christian Grefkes. 2011. “Dynamic Causal Modeling of Cortical Activity from the Acute to the Chronic Stage After Stroke.” Neuroimage 55 (3): 1147–58.

Reid, A. T., Headley, D. B., Mill, R. D., Sanchez-Romero, R., Uddin, L. Q., Marinazzo, D., … & Cole, M. W. (2019). Advancing functional connectivity research from association to causation. Nature neuroscience, 22(11), 1751–1760.

Riley, Jeff D, Vu Le, Lucy Der-Yeghiaian, Jill See, Jennifer M Newton, Nick S Ward, and Steven C Cramer. 2011. “Anatomy of Stroke Injury Predicts Gains from Therapy.” Stroke 42 (2): 421–26.

Schoffelen, Jan-Mathijs, and Joachim Gross. 2009. “Source Connectivity Analysis with MEG and EEG.” Human Brain Mapping 30 (6): 1857–65.

See, J., Dodakian, L., Chou, C., Chan, V., McKenzie, A., Reinkensmeyer, D. J., and Cramer, S. C. (2013). A standardized approach to the fugl-meyer assessment and its implications for clinical trials. Neurorehabilitation and neural repair, 27(8):732–741.

Schneider, M., Broggini, A. C., Dann, B., Tzanou, A., Uran, C., Sheshadri, S., … & Vinck, M. (2021). A mechanism for inter-areal coherence through communication based on connectivity and oscillatory power. Neuron, 109(24), 4050–4067.

Siems, Marcus, and Markus Siegel. 2018. “Dissociated Cortical Phase-and Amplitude-Coupling Patterns in the Human Brain.” bioRxiv, 485599.

Smith, Stephen M, Mark Jenkinson, Mark W Woolrich, Christian F Beckmann, Timothy EJ Behrens, Heidi Johansen-Berg, Peter R Bannister, et al. 2004. “Advances in Functional and Structural MR Image Analysis and Implementation as FSL.” Neuroimage 23: S208–19.

Srinivasan, Ramesh, William R Winter, Jian Ding, and Paul L Nunez. 2007. “EEG and MEG Coherence: Measures of Functional Connectivity at Distinct Spatial Scales of Neocortical Dynamics.” Journal of Neuroscience Methods 166 (1): 41–52.

Stam, Cornelis J. 2014. “Modern Network Science of Neurological Disorders.” Nature Reviews Neuroscience 15 (10): 683.

Takeuchi, Naoyuki, and Shin-Ichi Izumi. 2012. “Maladaptive Plasticity for Motor Recovery After Stroke: Mechanisms and Approaches.” Neural Plasticity 2012 (1): 359728.

Van Essen David C, Stephen M Smith, Deanna M Barch, Timothy EJ Behrens, Essa Yacoub, Kamil Ugurbil, Wu-Minn HCP Consortium, et al. 2013. “The WU-Minn Human Connectome Project: An Overview.” Neuroimage 80: 62–79.

Veerbeek Janne M, Gert Kwakkel, Erwin EH van Wegen, Johannes CF Ket, and Martijn W Heymans. 2011. “Early Prediction of Outcome of Activities of Daily Living After Stroke: A Systematic Review.” Stroke 42 (5): 1482–88.

Wang, Liang, Chunshui Yu, Hai Chen, Wen Qin, Yong He, Fengmei Fan, Yujin Zhang, et al. 2010. “Dynamic Functional Reorganization of the Motor Execution Network After Stroke.” Brain 133 (4): 1224–38.

Wang, Xiaomei, XiaoOu Mao, Lin Xie, Fen Sun, David A Greenberg, and Kunlin Jin. 2012. “Conditional Depletion of Neurogenesis Inhibits Long-Term Recovery After Experimental Stroke in Mice.” PloS One 7 (6): e38932.

Wanni Arachchige, Pradeepa Ruwan, Sadhani Karunarathna, Abdul Chalik Meidian, Ryo Ueda, Wataru Uchida, Masahiro Abo, and Atsushi Senoo. 2021. “Structural Connectivity Changes in the Motor Execution Network After Stroke Rehabilitation.” Restorative Neurology and Neuroscience 39 (4): 237–45.

Wieloch, Tadeusz, and Karoly Nikolich. 2006. “Mechanisms of Neural Plasticity Following Brain Injury.” Current Opinion in Neurobiology 16 (3): 258–64.

Wodeyar Anirudh. 2019. “Linking Structure to Function in Resting State Macroscale Neural Activity.” PhD thesis, UC Irvine.

Wodeyar, Anirudh, and Ramesh Srinivasan. 2022. “Structural Connectome Constrained Graphical Lasso for MEG Partial Coherence.” Network Neuroscience 6 (4): 1219–42.

Wodeyar, Anirudh, Jessica M. Cassidy, Steven C. Cramer, and Ramesh Srinivasan. “Damage to the structural connectome reflected in resting-state fMRI functional connectivity.” Network Neuroscience 4, no. 4 (2020): 1197–1218.

Woolrich, Mark W, Saad Jbabdi, Brian Patenaude, Michael Chappell, Salima Makni, Timothy Behrens, Christian Beckmann, Mark Jenkinson, and Stephen M Smith. 2009. “Bayesian Analysis of Neuroimaging Data in FSL.” Neuroimage 45 (1): S173–86.

Woytowicz, Elizabeth J, Jeremy C Rietschel, Ronald N Goodman, Susan S Conroy, John D Sorkin, Jill Whitall, and Sandy McCombe Waller. 2017. “Determining Levels of Upper Extremity Movement Impairment by Applying a Cluster Analysis to the Fugl-Meyer Assessment of the Upper Extremity in Chronic Stroke.” Archives of Physical Medicine and Rehabilitation 98 (3): 456–62.

Wu, Jennifer, Erin Burke Quinlan, Lucy Dodakian, Alison McKenzie, Nikhita Kathuria, Robert J Zhou, Renee Augsburger, et al. 2015. “Connectivity Measures Are Robust Biomarkers of Cortical Function and Plasticity After Stroke.” Brain 138 (8): 2359–69.

Yeh, Fang-Cheng, Sandip Panesar, David Fernandes, Antonio Meola, Masanori Yoshino, Juan C Fernandez-Miranda, Jean M Vettel, and Timothy Verstynen. 2018. “Population-Averaged Atlas of the Macroscale Human Structural Connectome and Its Network Topology.” Neuroimage 178: 57–68.

Zhou, R. J., Hondori, H. M., Khademi, M., Cassidy, J. M., Wu, K. M., Yang, D. Z., Kathuria, N., Erani, F. R., Dodakian, L., McKenzie, A., et al. (2018). “Predicting gains with visuospatial training after stroke using an eeg measure of frontoparietal circuit function.” Frontiers in neurology, 9:597

